# Genomic alterations in histologically clear low grade dysplasia resection margins forecast advanced neoplasia in ulcerative colitis

**DOI:** 10.64898/2026.09.10.26362759

**Authors:** Jennifer Fisher, Konstantin Bräutigam, Heather E Grant, Maximilian Mossner, Chirine Sakr, Ibrahim Al Bakir, Brian Saunders, Noriko Suzuki, Ana Wilson, Ann-Marie Baker, Ailsa Hart, Trevor A Graham

**Author notes:** Joint 1^st^ authors (Jennifer Fisher, Konstantin Bräutigam). co-senior authors (Ann-Marie Baker, Ailsa Hart, Trevor A Graham).

## Abstract

**Background:** Endoscopic resection is used for the management of dysplasia in ulcerative colitis (UC), but a subset of patients develop subsequent advanced neoplasia (AN), i.e., high-grade dysplasia and/or colorectal cancer, despite apparently complete resection with histologically clear margins. Current risk stratification relies predominantly on histopathological and procedural features, which does not capture underlying genomic field cancerization of morphologically normal mucosa.

**Objective:** To determine whether genomic alterations within histologically non-dysplastic resection margins are associated with subsequent neoplastic progression following endoscopic resection of UC-associated dysplasia.

**Design:** We analysed a retrospective cohort of 51 UC patients, who underwent endoscopic resection of dysplastic lesions at a tertiary referral centre, with a median follow-up of 1947 days. 11 of these had local progression. We performed low-coverage whole genome sequencing (lcWGS) of 56 dysplastic lesions and 91 adjacent histologically non-dysplastic margins. Somatic copy number alteration (CNA) burden and phylogenetic analysis were correlated with progression to AN.

**Results:** Histologically non-dysplastic margins from lesions that subsequently progressed to AN demonstrated significantly higher CNA burden compared with non-progressors. In multivariate analysis high margin CNA burden was independently associated with local progression, whereas histologically clear resection margins and *en bloc* excisions were not.

**Conclusion:** Histological clearance does not necessarily indicate clearance of the cancerised field in UC-associated dysplasia. Genomic detection of field cancerisation within histologically non-dysplastic resection margins identifies patients at increased risk of subsequent neoplasia and provides a potential biomarker for biologically informed post-resection surveillance.

**What is already known on this topic:**

- Endoscopic resection is increasingly used for colitis-associated dysplasia, but some patients subsequently develop advanced neoplasia despite histologically complete resections with clear margins.
- Current risk stratification post-resection relies on histopathological and procedural features, which likely do not capture the cancerised field.

**What this study adds:**

- Histomorphologically non-dysplastic margins can harbour genomic alterations that are shared with the dysplastic lesion, indicating a cancerised field.
- A high burden of copy number alterations in resection margins independently predicts subsequent progression to advanced neoplasia, beyond conventional measures of resection completeness.

**How this study might affect research, practice or policy:**

- Beyond histopathology, genomic assessment of resection margins is low-cost, fast and reproducible and may enable biologically informed risk stratification following endoscopic resection of colitis-associated dysplasia.

## Introduction

Colitis-associated colorectal cancer arises through a multistep evolutionary process in which chronic inflammation promotes the emergence, selection and expansion of genetically altered epithelial clones. Epithelial dysplasia is the well-recognized precursor within this process and remains the key target for surveillance and endoscopic intervention strategies in patients with longstanding inflammation in inflammatory bowel disease (IBD) (1–3). Historically, the detection of colorectal dysplasia frequently prompted colectomy because it was regarded as a marker of widespread neoplastic potential, reflecting an increased risk of synchronous and metachronous lesions and progression to invasive cancer (4,5). However, advances in surveillance, including the adoption of high-definition endoscopy and chromoendoscopy together with the development of advanced endoscopic resection techniques, have transformed the management of dysplasia in IBD (6). International guidelines, including recent updates to the British Society of Gastroenterology (BSG) guidelines and the American Gastroenterological Association (AGA) Clinical Practice Update, recommend endoscopic resection as the preferred management strategy for visible dysplastic lesions amenable to complete excision, offering patients an organ-preserving approach (3,7). Techniques including endoscopic mucosal resection (EMR) and endoscopic submucosal dissection (ESD) enable complete resection of appropriately selected visible dysplastic lesions, allowing many patients to avoid the morbidity and functional consequences associated with colectomy (8,9).

Despite these advances, risk stratification after endoscopic resection of IBD-associated dysplasia remains largely dependent on conventional clinicopathological parameters, including dysplasia grade, lesion morphology and resection margin status (7,9,10). *En bloc* excision with histologically negative margins (R0 resection) is generally interpreted as evidence of complete lesion clearance and is associated with a lower risk of local recurrence and subsequent advanced neoplasia (AN) (9,11). Conversely, incomplete excision, piecemeal resection, or positive margins are associated with an increased risk of residual or recurrent dysplasia (10,12). Nevertheless, a clinically important subset of patients develop recurrent AN despite apparently complete endoscopic resection with histologically clear margins (8,12). This suggests that histological assessment of the resected lesion and its immediate margins may not fully capture the underlying biological risk of future neoplastic development (13,14).

One explanation is that colitis-associated colorectal carcinogenesis is not solely a lesion-based process but occurs within a chronically inflamed and genetically altered “field” capable of accumulating widespread genomic alterations (1,2,13,15). Persistent inflammatory stress may drive the emergence, selection and expansion of genetically altered epithelial clones across the colonic mucosa, resulting in a field of genetically abnormal but morphologically non-neoplastic epithelium. This concept of “field cancerization” suggests that histologically non-dysplastic tissue may harbour genomic alterations that can subsequently progress to neoplasia (2,13,14). Consequently, a resection margin classified as histologically non-dysplastic may not necessarily represent complete eradication of the underlying cancerised field.

Recent genomic studies have begun to redefine the biological basis of neoplastic progression in IBD, demonstrating that genomic instability and clonal expansion can precede overt neoplasia and identify high risk patients (15–17). Our previous work demonstrated that the burden of somatic copy number alterations (CNAs) in low grade dysplasia predicts progression to AN beyond conventional clinicopathological features (17). These findings suggest that genomic alterations may provide clinically meaningful prognostication beyond histology alone.

However, genomic risk assessment has thus far remained largely lesion-centric, focusing on the dysplastic epithelium itself as the source of future risk. Whether histologically non-dysplastic colorectal mucosa surrounding resected lesions harbours measurable genomic alterations, and whether these alterations contribute to subsequent neoplastic progression, remains unknown. This question is clinically important because the tissue remaining after resection, rather than the excised lesion itself, might ultimately determine future cancer risk. In this study, we examine both dysplastic lesions and matched histomorphologically non-dysplastic resection margins using low coverage whole genome sequencing (lcWGS). By integrating lesion and margin genomic profiling, we aim to redefine the concept of complete excision from a purely histopathological endpoint towards a combined biologically informed measure of genomic clearance.

## Results

### Cohort characteristics and study design

We analyzed a retrospective cohort of 51 patients with ulcerative colitis who underwent endoscopic resection of dysplastic lesions at St Mark’s Hospital (London, UK). During a median follow-up of 1,947 days (IQR 1186 - 3220), 13 patients developed subsequent AN (high-grade dysplasia (HGD) or CRC). Of these, 11 patients developed HGD/CRC at the site of the index endoscopic resection (local progression) and were classified as progressors (P) for all subsequent analysis (**Figure 1**). Two patients developed HGD/CRC in a different bowel segment to the resected dysplasia, and were therefore not classified as progressors, as the primary outcome of this study was local progression following endoscopic resection.

**Figure 1.**
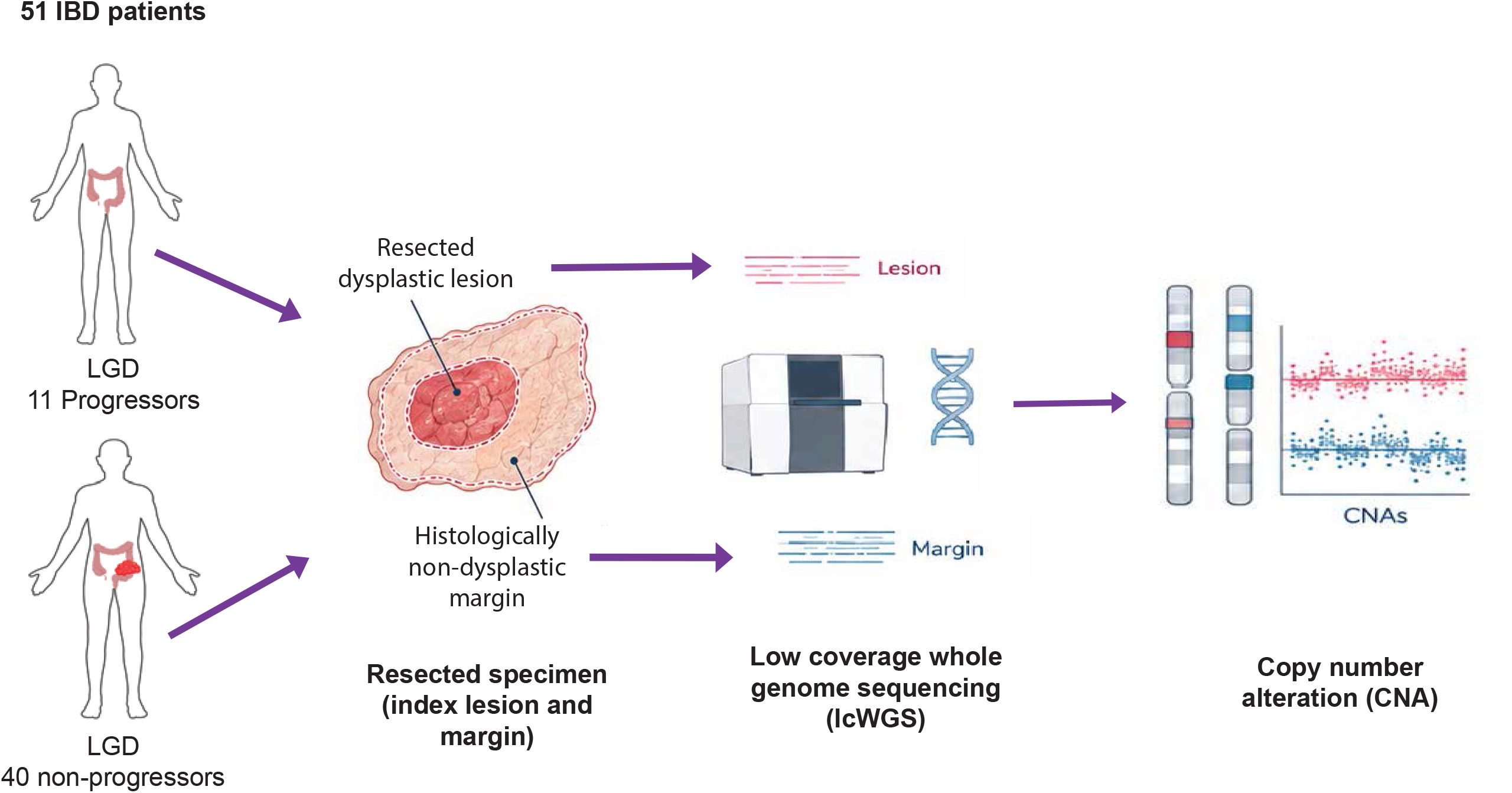
Study design and workflow. A total of 51 patients with inflammatory bowel disease (IBD)/ulcerative colitis (UC) were included in the study. Of these patients, 11 had low-grade dysplastic (LGD) lesions that progressed to advanced neoplasia, while the remaining 40 had LGD lesions that did not progress. The LGD index lesion and the margins were both histopathologically annotated, dissected, and subjected to low-coverage whole genome sequencing (lcWGS). Copy number alterations (CNAs) were quantified for all lesions and margins.

We analysed 162 spatially distinct tissue regions, representing 56 dysplastic lesions. This included 71 dysplastic regions and 91 regions of histologically non-dysplastic margins. 12 lesions subsequently developed HGD/CRC within the same bowel segment as the original resection, while 44 did not. 5 patients had two distinct dysplastic lesions, resected at the same time point. In some of these patients, local progression occurred at one index resection site but not the other, resulting in both progressing and non-progressing lesions for some individuals. The remaining 46 patients had a single LGD lesion (**Table 1**).

**Table 1.** Patient Characteristics.

| **Table 1. Patient Characteristics** |  |  |  |
| --- | --- | --- | --- |
|  | Local progressor | No local progression | Significance |
| Number of patients | 11 | 42 | NA |
| Number of lesions | 12 | 44 | NA |
| Resection technique |  |  | P = 0.736 (Fisher's exact test) |
| EMR | 18 | 6 |  |
| ESD | 22 | 5 |  |
| Number of margin regions sampled (median) | 3 | 2 | P = 0.382 (Wilcoxon rank sum) |
| Days to censor | 499 (IQR 308-814) | 2555 (IQR 1585 – 3728) | NA |
| Gender (% male) | 36.4 | 52.5 | P = 0.720 (fishers exact test) |
| Median age (years) | 62.5 (34-75) | 64 (27-84) | P = 0.442 (Wilcoxon rank rum) |
| Median duration of disease (years) | 26 (11-38) | 27.5 (1-52) | P = 0.9156 (Wilcoxon rank sum) |
| Proportion with PSC (%) | 18.2 | 2.4 | 0.106 (Fisher's exact test) |
| Spatial distribution | 69.2% left colorectum<br>30.8% right colon | 69.8% left colorectum<br>30.2% right colon | P = 1.00 (Fisher's exact test) |
| Lesion size (mm) | 14 (6-50) | 17 (5-65) | P = 0.811 Wilcoxon rank sum) |

### Histologically non-dysplastic margins of progressor lesions can harbour genomically altered clonal fields

We separately microdissected dysplasia and multiple regions of adjacent non-dysplastic “normal margin” tissue and on each region, we performed lcWGS, followed by genome-wide CNA profiling (**Figure 1**). Illustrative cases of a progressor and a non-progressor lesion are shown in **Figure 2**.

**Figure 2.**
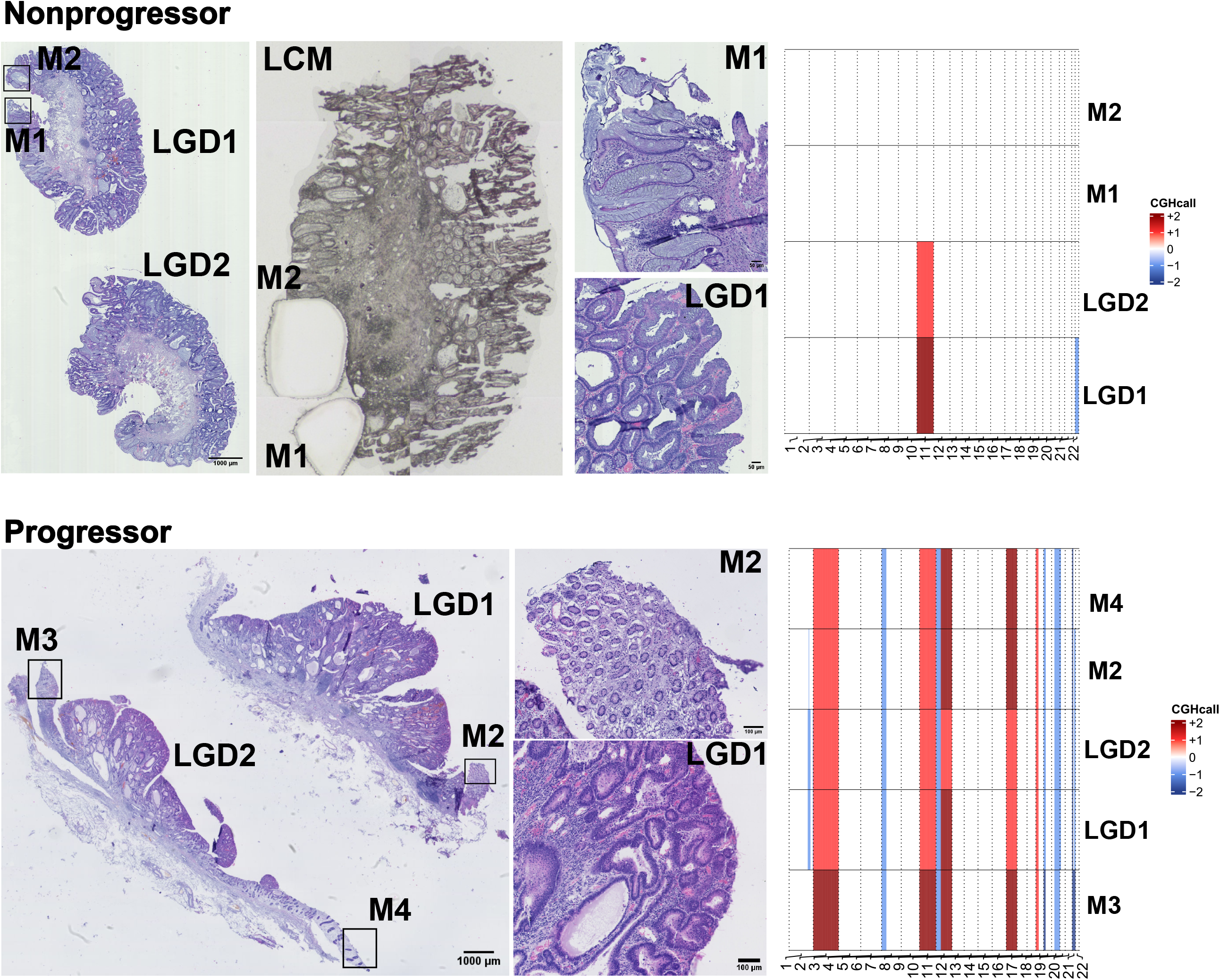
Field cancerisation in progressor lesions. Genomic profiling of low-grade dysplasia (LGD) lesions show aberrant copy numbers (heatmaps on right). However, non-progressor margins (M, M1 and M2) (top panel, high magnification shows representative histology of a histologically clear margin “M1” with minimal inflammation) remain largely genomically distinct, suggesting minimal field expansion. Progressor patients (lower panel) harbor clonal alterations extending into the surrounding histologically clear margins (representative margin “M2” middle panel at the bottom), which is consistent with field cancerisation (bottom right). Representative figure of laser microdissection (LCM) of margins “M1” and “M2” of a non-progressor (upper panel). Scale bars as indicated.

We analysed the CNA burden per lesion or margin; for multi-region sampled lesions and margins we used the sample with the highest CNA burden to capture the greatest extent of genomic alteration present within each lesion and to avoid dilution of focal high-burden regions. Progressor lesions had substantially greater burden of genomic alterations than non-progressors (21 [IQR 14.15-31.00) versus 6.5 altered segments [IQR 1.25-17.25], p = 0.011 by Wilcoxon rank-sum test, **Figure 3A,B**) This finding is consistent with previous observations that increasing genomic complexity within dysplastic tissue is associated with a greater likelihood of progression, and it held true if genomic alterations were instead quantified by percentage genome altered (PGA; median 23.6% vs 4.8%, p = 0.005).

**Figure 3:**
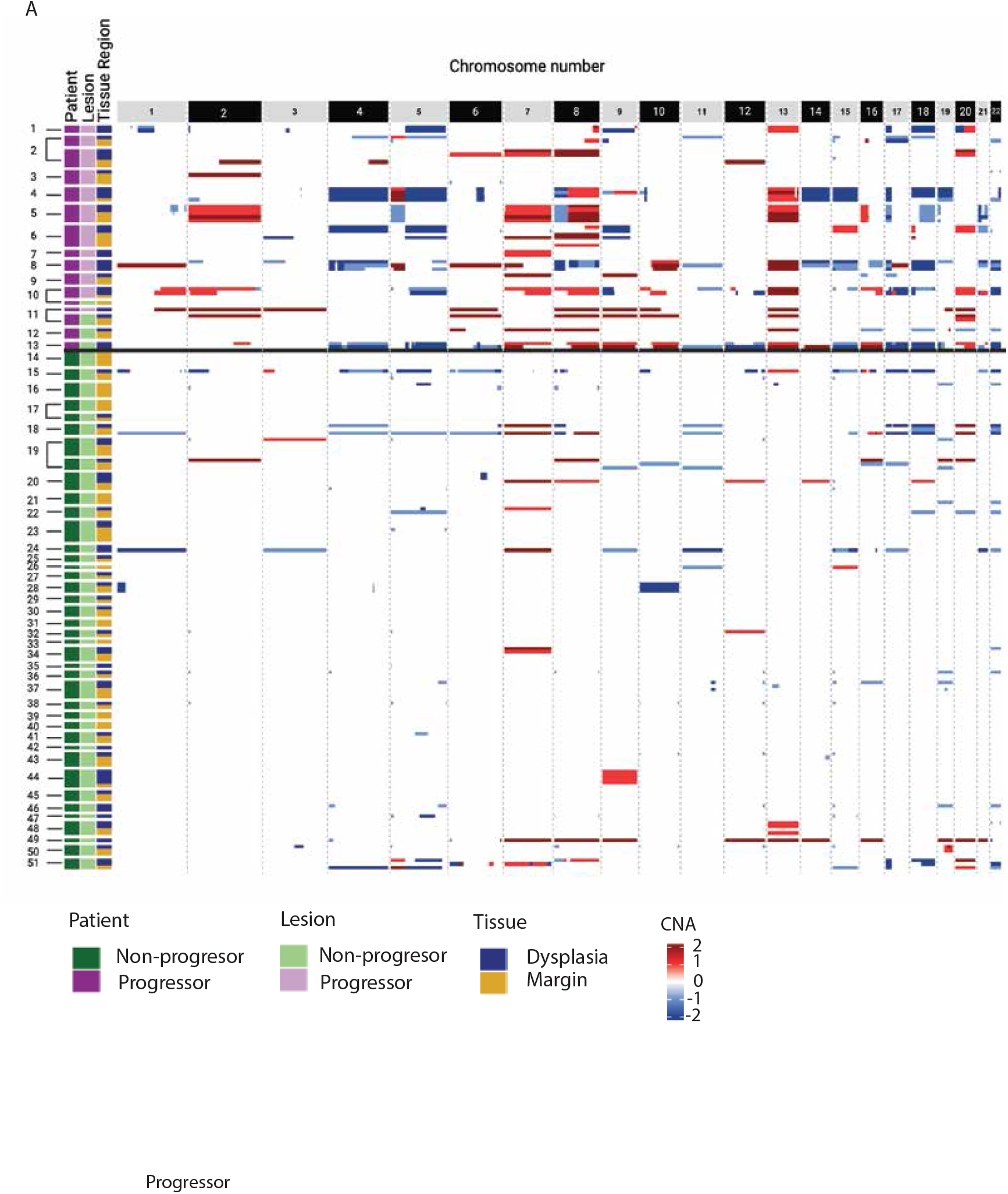
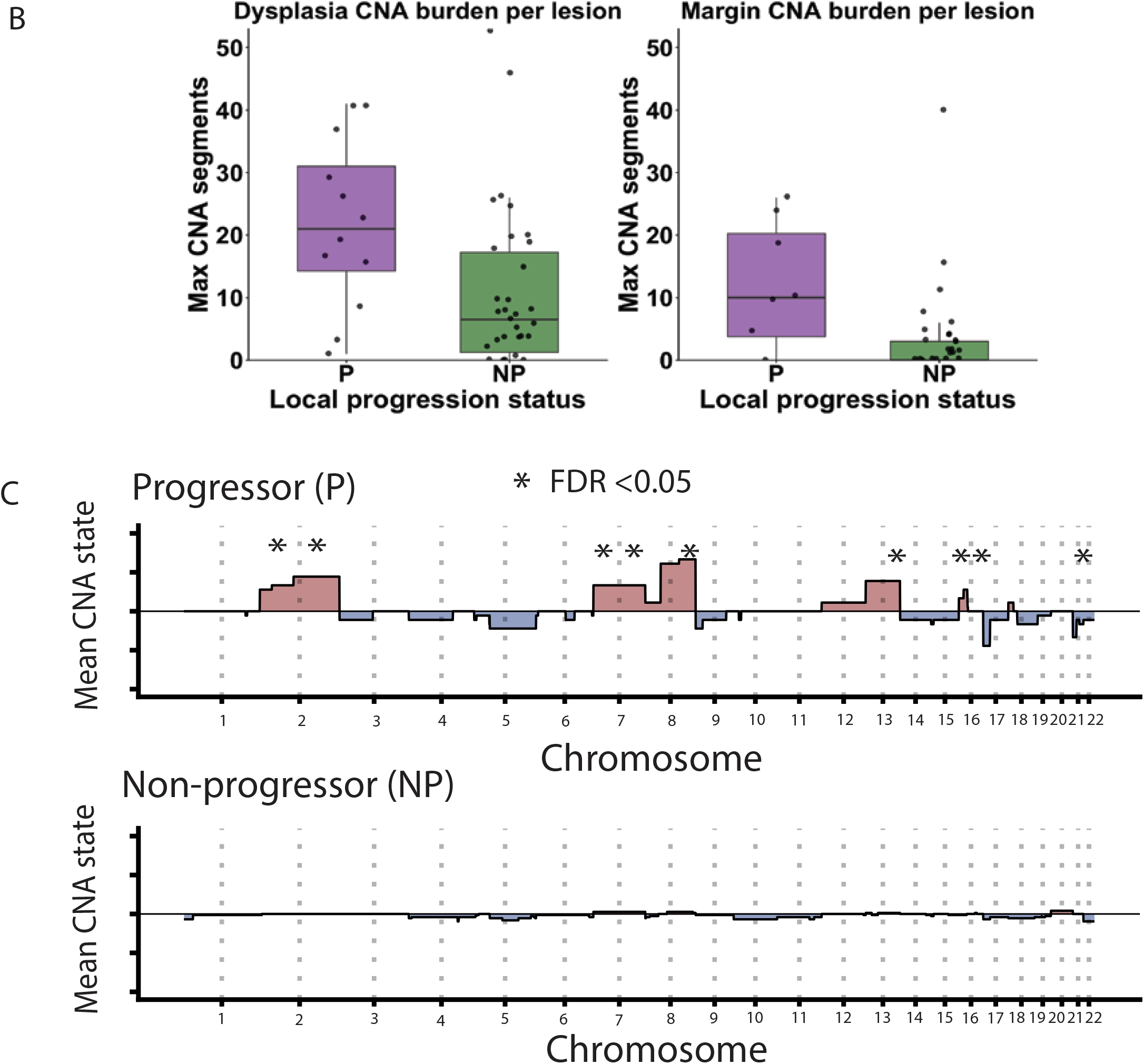
Chromosomal copy number alteration (CNA) burden by local progression status and tissue region. (A) Heatmap of genome-wide CNA calls across all samples, ordered by patient status, lesion and tissue region (dysplasia vs margin). Samples are stratified by progression status. (B) Lesion level CNA burden measured as the maximum number of CNA segments altered per lesion. (C) Chromosome wide CNA profiles comparing margin tissue from progressor and non-progressor patients.

Histologically non-dysplastic margins from progressors also demonstrated a substantially higher CNA burden (median 10 [IQR 3.75-20.75] vs 0 [IQR 0-3] altered segments; p = 0.010 by Wilcoxon rank-sum test, Figure **3B**) and PGA (median 10.5% vs 0.0%, p = 0.011). We noted that progressor dysplastic lesions and margin samples shared numerous ancestral copy number events, indicating that both arose from a common clonal precursor and that the neoplastic field extended beyond the histological boundary of the lesion. In contrast, non-progressors demonstrated relatively few genomic alterations, with limited clonal sharing between dysplastic and adjacent non-dysplastic margins. These findings demonstrate that “histologically clear” resection margins can differ in their underlying genomic state, with some harbouring genetically altered colorectal mucosa that is not detectable by routine pathological assessment.

The average genome-wide CNA profiles revealed distinct patterns of chromosomal alterations between progressors and non-progressor margins, with recurrent CNA events enriched in progressors (**Figure 3C**). Differential chromosome-arm level analysis identified enrichment of several recurrent CNA events in progressor margins, including 8q gain (p <0.001 by Fisher’s exact test with FDR correction) and 17p loss (p = 0.034 by Fisher’s exact test with FDR correction). Notably these regions encompass key cancer-associated loci including *MYC* at 8q24 and *TP53* at 17p13. These findings indicate that histologically non-dysplastic margins from progressor lesions retain measurable genomic alterations, in contrast to the largely genomically quiet margins observed in non-progressors.

### Margin CNA burden predicts progression following endoscopic resection

Given that histologically non-dysplastic margins from progressor lesions demonstrated increased CNA burden, we next investigated whether this could be used to identify lesions at increased risk of subsequent local progression following endoscopic resection.

Margin CNA burden was moderately correlated with lesion CNA burden (Spearman’s ⍰= 0.51, p = 0.002), although several lesions demonstrated discordant lesion and margin CNA burdens (**Figure 4A**).

**Figure 4:**
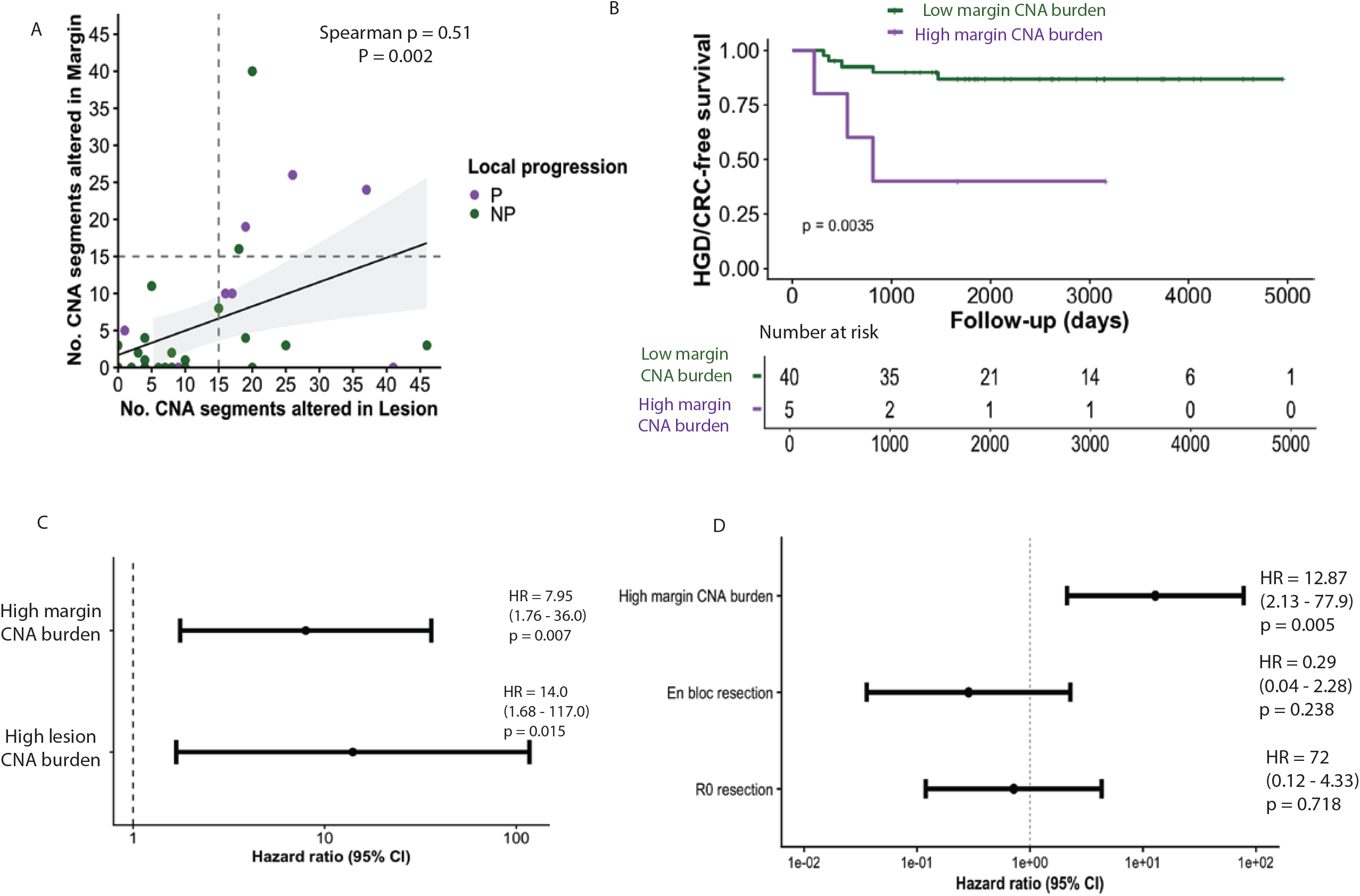
Genomic alterations within histologically non-dysplastic resection margins predict subsequent neoplastic progression. (A) Scatter plot demonstrating the correlation between dysplastic lesion and margin CNA burden. (B) Kaplan-Meier analysis demonstrating HGD/CRC-free survival stratified by margin somatic copy number alteration (CNA) burden. (C) Univariate Cox proportional hazards analysis evaluating the association between margin CNA burden and dysplastic lesion CNA burden with subsequent local progression. (D) Multivariable Cox proportional hazards evaluating the association between margin CNA burden and subsequent progression, adjusting for conventional endoscopic and histopathological parameters.

Lesions were stratified by margin CNA burden using a previously reported threshold of >14.5 altered CNA segments to define high risk lesions (17). Margins with >14.5 altered CNA segments were classified as having high margin CNA burden, while those with ≤ 14.5 altered CNA segments were classified as having low margin CNA burden.

High margin CNA burden was associated with significantly reduced progression-free survival (log rank p = 0.0035) (**Figure 4B**). In other words, a high CNA burden within histologically non-dysplastic mucosa identified lesions at increased risk of progression to AN following endoscopic resection.

Given the observed correlation between lesion and margin CNA burden, univariable Cox proportional hazards models were constructed to compare the prognostic performance of margin CNA burden and dysplastic lesion CNA burden (**Figure 4C**). High margin CNA burden was associated with an increased risk of subsequent progression (HR 7.95, 95% CI 1.76-36.0, p = 0.007). High lesion CNA burden was also associated with progression risk, performing similarly to our previous study (17) (HR 14.0, 95% CI 1.68-117, p = 0.015).

Multivariable modeling was performed to determine whether margin CNA burden provided prognostic information beyond established procedural factors (**Figure 4D**). High margin CNA burden remained independently associated with subsequent local progression (adjusted HR 12.87, 95% CI 2.13 – 77.90, p =0.005). In contrast, neither histologically clear margins, R0 status, (HR 0.72 (95% CI 0.12-4.33, p = 0.718) nor *en bloc* resection (HR 0.29, 95% CI 0.04-2.28, p = 0.238) were independently associated with progression risk. These findings suggest that genomic alterations in resection margins (i.e. a cancerised field) provides prognostic information beyond conventional measures of endoscopic resection and completeness.

### Phylogenetic analysis reveals spatially extended clonal fields spanning dysplastic lesions and histologically normal margins in progressors

Building upon our observation of clonal sharing of CNAs between margins and lesions (**Figure 2**), we performed CNA-based phylogenetic analysis for each patient. Using phylogenetic trees and matched CNA heatmaps we demonstrated distinct patterns of clonal evolution according to clinical outcome (**Figure 5A**). Dysplastic and histologically non-dysplastic margin samples shared multiple early CNA events, with a common ancestral trunk before diverging into spatially distinct subclones, consistent with the presence of an evolutionarily advanced clonal field extending beyond the visible dysplastic lesion. In contrast, lesion and margin samples from the representative non-progressor showed relatively simple phylogenetic architectures with limited shared ancestry and less extensive branching, consistent with restricted clonal expansion.

**Figure 5:**
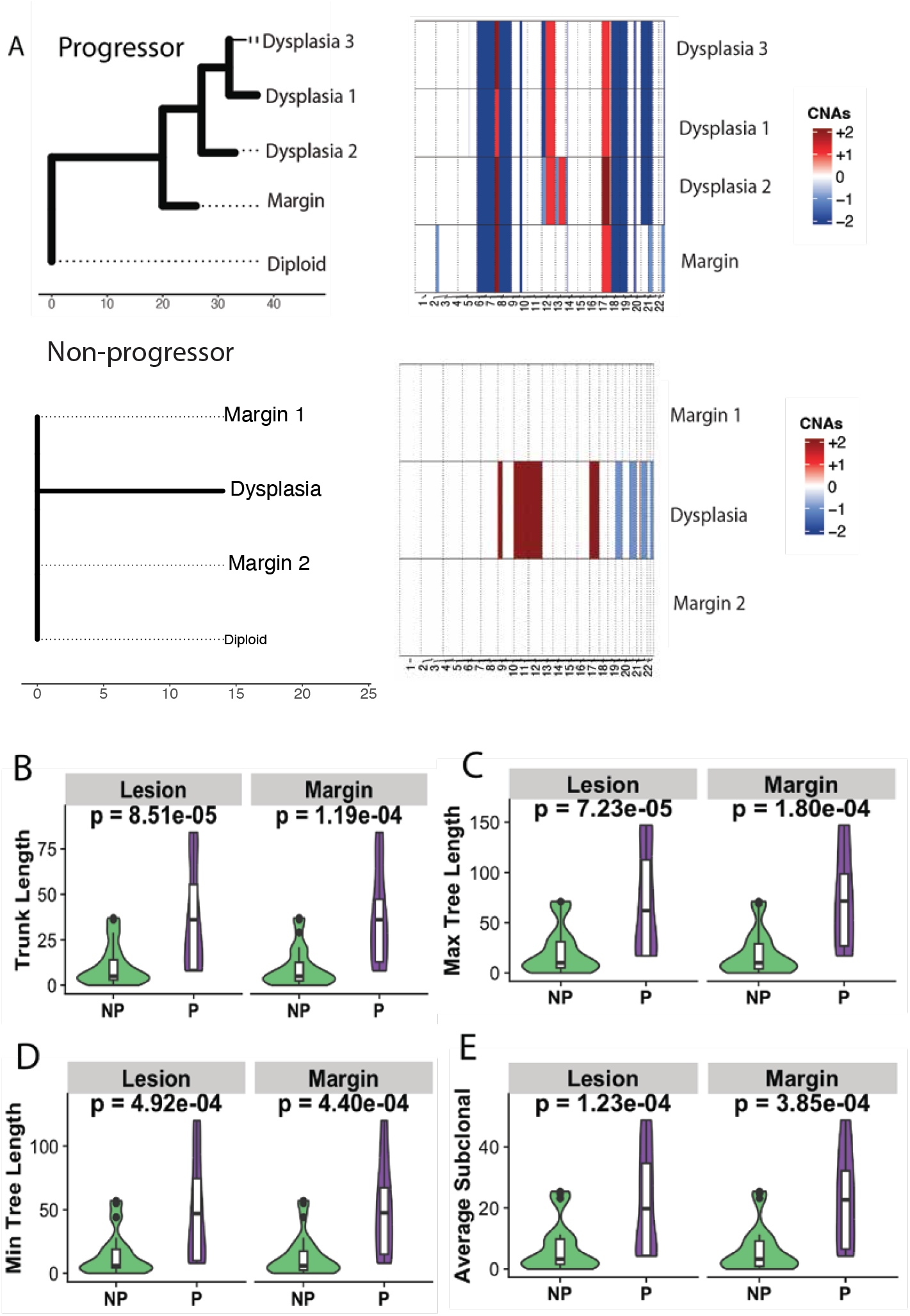
Phylogenetic analysis of lesion and margin regions. (A) Representative phylogenetic trees and matched copy number alteration (CNA) profiles from a progressor and non-progressor case. (B-E) Comparison of phylogenetic tree characteristics between non-progressor (NP) and progressor (P) lesions and their matched resection margins: (B) trunk length, shared clonal CNA events; (C) maximum tree length; (D) minimum tree length; and (E) average subclonal distance, representing the mean evolutionary distance from the trunk to terminal bran ches.

### Most copy number alterations are early and clonal

Quantitative phylogenetic analysis confirmed significantly greater evolutionary diversification in progressors across both dysplastic lesions and histologically non-dysplastic margins (**Figure 5B-E**). Within dysplastic regions, progressors had significantly longer ancestral trunks (representing shared copy-number evolution before subclonal divergence; median 36.0 vs 5.0), greater maximum tree length (the number of inferred copy-events to the of the most highly altered region; 62.0 vs 10.0), greater minimum tree length (events to the least altered region; 47.0 vs 6.0) and increased average subclonal distance (mean evolutionary distance from the trunk to the terminal branches; 19.8 vs 3.3) compared with non-progressors (all p < 0.001, Wilcoxon rank-sum test). These findings reflect prolonged clonal evolution before diversification together with substantially greater intralesional evolutionary heterogeneity in lesions that subsequently progressed. Similar differences were observed in histologically non-dysplastic margins, with progressors showing greater maximum tree length (median 71.5 vs 10.0, p <0.001), minimum tree length (47.5 vs 6.0, p = 0.003), trunk length (36.0 vs 5.0, p <0.001) and average subclonal distance (22.6 vs 3.3, p = 0.001). Thus, the evolutionary features associated with progression within dysplastic lesions were also evident in histologically non-dysplastic margins, supporting the presence of a genetically altered field extending beyond the morphologically apparent lesion.

Notably, the phylogenetic proximity between dysplastic lesions and their corresponding histologically normal margins suggests that, once established, the clonal field closely mirrors that of the visible dysplastic lesion. These findings indicate that in progressors, the dysplastic lesion represents only the morphologically apparent component of a substantially larger, evolutionarily mature field of genetically altered mucosa. This demonstrates that histologically normal resection margins from progressors represent genetically advanced tissue rather than uninvolved mucosa, providing evolutionary evidence for field cancerization in UC-associated neoplasia.

## Discussion

A major challenge in UC-associated dysplasia is the limited ability of conventional histopathology to accurately predict which patients will develop subsequent neoplasia following apparently complete endoscopic resection (5,6). In this study, we demonstrate that histomorphologically non-dysplastic resection margins frequently harbour substantial genomic alterations associated with future neoplastic progression. Margin CNA burden emerged as the strongest independent predictor of progression and provided prognostic information beyond conventional procedural and pathological parameters including R0 resection status and *en bloc* excision. These findings suggest that histological clearance and biological clearance are not synonymous.

Phylogenetic analysis demonstrated that, in progressor cases, dysplastic lesions and adjacent non-dysplastic margins frequently shared early ancestral CNA events, consistent with emergence from a common genetically altered clone. Increased phylogenetic complexity was observed not only within dysplastic lesions but also within surrounding margin regions, indicating that evolutionary diversification extends beyond the visible dysplastic lesion. Together, these findings support a model of UC-associated neoplasia driven by spatially extended clonal fields encompassing both dysplastic epithelium and histologically non-dysplastic mucosa (2). In contrast, non-progressors generally exhibited more restricted evolutionary relationships between lesion and margin, consistent with either limited field involvement or containment of neoplastic evolution within a more localized clone.

Importantly, our findings suggest that progression risk is determined not solely by the biology of the index lesion but by the extent and persistence of the surrounding clonal field. Notably, a subset of non-progressor cases also demonstrated elevated CNA burden within resection margins. This suggests that absence of progression does not necessarily indicate biologically low-risk disease. One possibility is that complete endoscopic resection removes the entirety of a genetically altered field, thereby interrupting a potential trajectory towards neoplastic progression. Alternatively, the time to progression may exceed the minimum 3 years of follow-up available in this study. An important consideration is that, particularly following piecemeal resection, a subsequent lesion arising at the index site cannot always be distinguished as residual disease from a new neoplastic event arising within the surrounding field. However, margin sampling captures the genomic state of surrounding mucosa at the time of resection, providing a baseline measure of field-level genomic alteration. Importantly, margin CNA burden remained independently associated with subsequent advanced neoplasia in multivariable analysis, supporting its potential value as a marker of underlying field risk beyond the characteristics of the index lesion and completeness of resection. Together, these findings suggest that progression may reflect both the biological potential of the field and whether genetically altered mucosa remains after resection.

These findings have important implications for post-resection surveillance and risk stratification. Current management strategies rely predominantly on histological assessment of resection margins and lesion characteristics, neither of which captures underlying field effects (4–6). Our findings suggest that future management could move beyond histological assessment alone and towards molecular assessment of the residual field. Margin CNA burden represents one such biomarker, but the broader implication is that defining the spatial extent of the genetically altered field may become as important as characterising the dysplastic lesion itself. Future approaches should therefore not only seek to measure genomic risk after resection but also to define the spatial extent of genetically altered mucosa during endoscopic assessment. Previous studies using DNA ploidy analysis and spatial molecular profiling have shown that genomic abnormalities extend beyond histologically dysplastic regions, supporting the feasibility of molecular field mapping. Integration of molecular biomarkers with advanced endoscopic imaging could ultimately enable real-time identification of genomically altered mucosa, allowing resection to be guided by the true extent of the neoplastic field rather than by morphological abnormalities alone (13,17,18). Prospective validation will be required to determine whether genomic assessment of resection margins can be incorporated into routine clinical practice and used to guide surveillance intensity following endoscopic resection.

In summary, we show that UC-associated dysplasia frequently arises within spatially extensive cancerised fields that are not captured by conventional histopathology. Progression is associated with persistent, clonally related fields spanning dysplastic lesions and adjacent non-dysplastic mucosa, and genomic assessment of these fields provides prognostic information beyond conventional pathological assessment. Together, these findings provide a link between field cancerization and post resection outcomes, supporting the development of molecularly informed approaches for risk stratification, endoscopic management and surveillance in UC-associated dysplasia.

## Methods

### Study design

We performed a retrospective cohort study of patients with ulcerative colitis treated at St Mark’s Hospital (London, UK). Patients were stratified by clinical outcome into progressors, defined as those who developed advanced neoplasia (high grade dysplasia or colorectal cancer) during follow up, and non-progressors, defined as patients without evidence of advanced neoplasia during a minimum follow-up of 3 years.

All patients were followed longitudinally within a clinical surveillance programme, with variable follow-up duration across individuals. Progression events occurred during follow up and were analyzed using time-to-event methods with right censoring at last clinical assessment. Clinical data were extracted from a curated database linked to histopathological and sequencing datasets.

### Histology and sample preparation

Formalin-fixed paraffin-embedded (FFPE) resections were sectioned at 10 μm, stained with haematoxylin and eosin (H&E) and scanned using a NanoZoomer digital slide scanner (Hamamatsu Photonics, Japan) for further review and annotation. All samples were digitized, reviewed and regions of interest annotated by a board-certified pathologist (KB), including dysplastic and adjacent non-dysplastic mucosa. Non-dysplastic tissue was separately dissected from the dysplastic lesion; in piecemeal resections lateral margins were dissected where clearly identifiable.

### DNA extraction and library preparation

Resection specimens with clearly identifiable histomorphologically ‘clear’ non-dysplastic resection margins, i.e. lateral to conventional low-grade dysplasia (LGD), underwent separate dissection of all margins and of the dysplastic lesion present on the slide. Three 10 µm sections per sample were dissected for DNA extraction. A total of 56 lesions were sequenced, of which 35 had matched lesion and margin data, 11 had dysplastic lesion data only, and 10 had margin data only.

The slides were dewaxed in xylene for 2 x 5 minutes and rehydrated using 5-minute incubations in ethanol dilutions. The margins were microdissected using laser microdissection (Leica LMD6, Leica Microsystems, Wetzlar, Germany) on PEN-membraned 10 µm sections. Larger lesions were needle macrodissected. DNA was then extracted using a modified protocol of the High Pure FFPET DNA Isolation Kit (Roche Life Science, Basel, Switzerland).

### Low coverage whole genome sequencing (lcWGS)

A minimum of 3.75 ng of input DNA was used for library preparation using the NEBNext Ultra II DNA Library Prep Kit for Illumina (New England Biolabs). The libraries were barcoded using the NEBNext Multiplex Oligos for Illumina kit and subjected to 16 cycles of library amplification. The final library quantity was confirmed using a Qubit fluorometer, and the quality of the libraries (fragment size distribution and the presence of unbound adapters) was verified using an Agilent High Sensitivity D1000 DNA TapeStation system. Libraries were sequenced if the peak fragment size was greater than 184 base pairs.

Samples were equimolar pooled and sequenced to a target 0.1× coverage using the Illumina NextSeq 500. After sequencing, we used the same bioinformatics pipeline as in (12). Briefly, reads were trimmed using Skewer v.0.2.2, then checked with fastQC v0.11.9, mapped to UCSC hg19 using BWA-MEM, keeping only reads with mapping Quality 37 or above, and duplicates removed with picard MarkDuplicates. BAM QC was performed using Qualimap BamQC (v2.2.2), and only samples with at least 1 million mapped reads were included in downstream analyses.

### Copy number alteration analysis

Genome-wide copy number profiles were generated from low-coverage sequencing data using a fixed genomic binning framework, following the approach previously described by Al Bakir et al (17). Briefly, sequencing read depth was quantified across predefined genomic bins and normalised to identify regions of relative copy-number gain or loss. The resulting copy-number profiles were segmented to identify contiguous regions showing deviation from the expected diploid baseline. CNA burden was quantified as the total number of CNA segments identifed across the genome, with higher values representing greater genomic instability.

Each lesion consisted of multiple spatial samples, and CNA burden was quantified at the sample level. Lesion level summaries were derived by aggregating sample-level measurements, and patient level variables were derived from lesion-level data.

### Phylogenetic analysis

To assess copy number evolution and intralesional heterogeneity, phylogenetic trees were reconstructed using MEDICC2 (19), for lesions with matched dysplastic and histologically non-dysplastic margin samples, following a previously described approach (17). Branch lengths represented the number of inferred copy-number events separating genomic states. Tree-derived measures were compared between progressor and non-progressor lesions using the Wilcoxon rank-sum test.

### Outcome definitions

The primary outcome was local progression, defined as the development of advanced neoplasia (high grade dysplasia or colorectal cancer) within the same bowel segment as the index lesion.

Primary analysis focused on local progression to ensure spatial concordance between the sequenced tissue and subsequent outcome. Neoplastic events occurring at anatomically distinct colorectal sites were not classified as local progression events for the primary analysis.

### Outcome analysis

Time-to-event analyses were performed on Kaplan–Meier survival curves, with differences between groups assessed using log-rank tests implemented with the R “survival” package. Multivariable Cox models were used to estimate hazard ratios (HRs) and 95% confidence intervals (CIs). Forest plots were generated using ggplot2 to visualize effect estimates and confidence intervals.

### Statistical analysis

Statistical analyses were performed in the R statistical computing environment (version 4.6.1) using RStudio. All p values were calculated using two-tailed tests, with statistical significance set at p < 0.05. Forest plots show the 95% confidence intervals (CIs) of hazard ratios (HRs) and the P values of the covariates were derived from Wald tests.

### Ethics approval

This was a retrospective anonymized analysis of archival FFPE patient samples. The study was approved by the UK REC approval number 18/LO/2051.

## Data Availability

All data produced in the present study are available upon reasonable request to the authors

## Data availability statement

Data are available from the corresponding authors upon reasonable request.

## Footnotes

### Contributors

Study conception and design: JF, KB, TAG, A-MB, ALH, AW. Bioinformatics: JF, HEG. Tissue processing and genomics: KB, JF, A-MB, CS, MM. Pathology and endoscopy data: JF, KB, NS, BS. Paper writing: JF, KB, A-MB, TAG, ALH.

## Acknowledgements

We thank the patients and their families for supporting research and donating samples for research. We thank the Genomics and Histopathology core facilities at the Institute of Cancer Research (ICR) for their support.

## Patient and public involvement

Patients and/or the public were not involved in the design, or conduct, or reporting, or dissemination plans of this research.

## References

1. Choi CHR, Bakir IA, Hart AL, Graham TA. Clonal evolution of colorectal cancer in IBD. Nat Rev Gastroenterol Hepatol. 2017 Apr;14(4):218–29. doi:10.1038/nrgastro.2017.1

2. Baker AM, Cross W, Curtius K, Bakir IA, Choi CHR, Davis HL, et al. Evolutionary history of human colitis-associated colorectal cancer. Gut. 2019 Jun 1;68(6):985–95. doi:10.1136/gutjnl-2018-316191 PubMed PMID: 29991641.

3. East JE, Gordon M, Nigam GB, Sinopoulou V, Bateman AC, Din S, et al. British Society of Gastroenterology guidelines on colorectal surveillance in inflammatory bowel disease. Gut. 2025 Apr 30. doi:10.1136/gutjnl-2025-335023 PubMed PMID: 40306978.

4. Farraye FA, Odze RD, Eaden J, Itzkowitz SH. AGA Technical Review on the Diagnosis and Management of Colorectal Neoplasia in Inflammatory Bowel Disease. Gastroenterology. 2010 Feb 1;138(2):746–774.e4. doi:10.1053/j.gastro.2009.12.035

5. Blackstone MO, Riddell RH, Rogers BH, Levin B. Dysplasia-associated lesion or mass (DALM) detected by colonoscopy in long-standing ulcerative colitis: an indication for colectomy. Gastroenterology. 1981 Feb;80(2):366–74. PubMed PMID: 7450425.

6. Maresca R, Calabrese G, Marchitto SA, Schepis T, Pecere S, Maida M, et al. Advanced diagnostic and resection endoscopic techniques in managing colitis-associated neoplasia: standard of care or still utopia? Best Pract Res Clin Gastroenterol. 2025 Sep 1;Cutting Edge Developments and Novel Targets in IBD78:102051. doi:10.1016/j.bpg.2025.102051

7. Murthy SK, Feuerstein JD, Nguyen GC, Velayos FS. AGA Clinical Practice Update on Endoscopic Surveillance and Management of Colorectal Dysplasia in Inflammatory Bowel Diseases: Expert Review. Gastroenterology. 2021 Sep 1;161(3):1043–1051.e4. doi:10.1053/j.gastro.2021.05.063

8. Alkandari A, Thayalasekaran S, Bhandari M, Przybysz A, Bugajski M, Bassett P, et al. Endoscopic Resections in Inflammatory Bowel Disease: A Multicentre European Outcomes Study. J Crohns Colitis. 2019 Oct 28;13(11):1394–400. doi:10.1093/ecco-jcc/jjz075 PubMed PMID: 30994915.

9. Mohapatra S, Sankaramangalam K, Lopimpisuth C, Moninuola O, Simons M, Nanavati J, et al. Advanced endoscopic resection for colorectal dysplasia in inflammatory bowel disease: a meta-analysis. Endosc Int Open. 2022 May;10(5):E593–601. doi:10.1055/a-1784-7063

10. Laine L, Kaltenbach T, Barkun A, McQuaid KR, Subramanian V, Soetikno R, et al. SCENIC international consensus statement on surveillance and management of dysplasia in inflammatory bowel disease. Gastrointest Endosc. 2015 Mar 1;81(3):489–501.e26. doi:10.1016/j.gie.2014.12.009

11. Malik TF, Sabesan V, Mohan BP, Rahman AU, Othman MO, Draganov PV, et al. Efficacy and safety of endoscopic submucosal dissection for colorectal dysplasia in patients with inflammatory bowel disease: a systematic review and meta-analysis. Clin Endosc. 2024 May;57(3):317–28. doi:10.5946/ce.2023.205 PubMed PMID: 38419168; PubMed Central PMCID: PMC11133987.

12. Mohan BP, Khan SR, Chandan S, Kassab LL, Ponnada S, Asokkumar R, et al. Endoscopic resection of colon dysplasia in patients with inflammatory bowel disease: a systematic review and meta-analysis. Gastrointest Endosc. 2021 Jan;93(1):59–67.e10. doi:10.1016/j.gie.2020.06.048 PubMed PMID: 32592777.

13. Leedham SJ, Graham TA, Oukrif D, McDonald SAC, Rodriguez–Justo M, Harrison RF, et al. Clonality, Founder Mutations, and Field Cancerization in Human Ulcerative Colitis–Associated Neoplasia. Gastroenterology. 2009 Feb 1;136(2):542–550.e6. doi:10.1053/j.gastro.2008.10.086

14. Galandiuk S, Rodriguez–Justo M, Jeffery R, Nicholson AM, Cheng Y, Oukrif D, et al. Field Cancerization in the Intestinal Epithelium of Patients With Crohn’s Ileocolitis. Gastroenterology. 2012 Apr 1;142(4):855–864.e8. doi:10.1053/j.gastro.2011.12.004 PubMed PMID: 22178590.

15. Lai LA, Risques RA, Bronner MP, Rabinovitch PS, Crispin D, Chen R, et al. Pan-colonic field defects are detected by CGH in the colons of UC patients with dysplasia/cancer. Cancer Lett. 2012 Jul 28;320(2):180–8. doi:10.1016/j.canlet.2012.02.031 PubMed PMID: 22387989; PubMed Central PMCID: PMC3406733.

16. Salk JJ, Salipante SJ, Risques RA, Crispin DA, Li L, Bronner MP, et al. Clonal expansions in ulcerative colitis identify patients with neoplasia. Proc Natl Acad Sci U S A. 2009 Dec 8;106(49):20871–6. doi:10.1073/pnas.0909428106 PubMed PMID: 19926851; PubMed Central PMCID: PMC2779829.

17. Bakir IA, Curtius K, Cresswell GD, Grant HE, Nasreddin N, Smith K, et al. Low-coverage whole genome sequencing of low-grade dysplasia strongly predicts advanced neoplasia risk in ulcerative colitis [Internet]. 2025 Jan 29. doi:10.1136/gutjnl-2024-333353

18. Levine DS, Rabinovitch PS, Haggitt RC, Blount PL, Dean PJ, Rubin CE, et al. Distribution of aneuploid cell populations in ulcerative colitis with dysplasia or cancer. Gastroenterology. 1991 Nov 1;101(5):1198–210. doi:10.1016/0016-5085(91)90068-V

19. Kaufmann TL, Petkovic M, Watkins TBK, Colliver EC, Laskina S, Thapa N, et al. MEDICC2: whole-genome doubling aware copy-number phylogenies for cancer evolution. Genome Biol. 2022 Nov 14;23(1):241. doi:10.1186/s13059-022-02794-9

